# Serum cFAS Content Correlates with Incidence of Peripheral Arterial Disease

**DOI:** 10.1101/2025.04.06.25325325

**Authors:** Wahid Abu-Amer, Khaled Shorbaji, Rodrigo Meade, Sophia R Pyeatte, Larisa Belaygorod, Mohamed S Zaghloul, Shahab Hafezi, Amanda Penrose, Batool Arif, Stephen G Wu, Clay F Semenkovich, Mohamed A Zayed

**Affiliations:** Washington University School of Medicine, Department of Surgery, Section of Vascular Surgery, St. Louis, MO, USA; Olin Business School, Washington University in St. Louis, MO, USA; Washington University School of Medicine, Department of Medicine, Division of Endocrinology, Metabolism and Lipid Research, St. Louis, MO, USA; Washington University School of Medicine, Department of Cell Biology and Physiology, St. Louis, MO, USA; Washington University School of Medicine, Department of Radiology, St. Louis, MO; Washington University School of Medicine, Division of Molecular Cell Biology, St. Louis, MO; Washington University School of Medicine, Division of Surgical Sciences, St. Louis, MO; Washington University, McKelvey School of Engineering, Department of Biomedical Engineering, St. Louis, MO; Veterans Affairs St. Louis Health Care System, St. Louis, MO

**Keywords:** Peripheral Arterial Disease, Serum Biomarker, Diabetes, Chronic Limb Threatening Ischemia

## Abstract

**Background:** No reliable serum diagnostic test currently exists for peripheral arterial disease (PAD). We previously observed that serum circulating Fatty Acid Synthase (cFAS) is elevated in individuals with chronic limb threatening ischemia (CLTI).

**Objectives:** We hypothesized that cFAS can be an independent diagnostic biomarker for PAD and CLTI.

**Methods:** Patients with/without PAD and CLTI were retrospectively reviewed. Total serum cFAS content was evaluated using ELISA and normalized to total protein. Patient demographics and PAD incidence were collected via chart review. Serum cFAS and demographics were compared, and regression analysis was used to determine the correlation between cFAS and PAD incidence, and the impact of co-morbidities on cFAS content.

**Results:** A total 347 patients met inclusion criteria. Of these, 34 were healthy controls without PAD (Group 1), 164 had PAD (Group 2), and 149 had CLTI (Group 3). Compared to Group 1, the remaining groups were significantly older, had more males, and had higher incidence of cardiovascular co-morbidities (p<0.001). Compared to Group 1, Groups 2 and 3 had significantly higher serum cFAS content (p=0.007). ROC analysis revealed an optimal cutoff of 340pg/mg protein for cFAS in distinguishing between individuals with or without PAD (p<0.001), and 490pg/mg protein in distinguishing between those with PAD and those with CLTI (p=0.015).

**Conclusions:** Our study demonstrates that cFAS is an independent serum-based diagnostic biomarker for PAD, can distinguish between patients with PAD versus CLTI, and may serve as a predictive variable for identifying patients with highest risk of disease progression.

**CONDENSED ABSTRACT:** There are currently no reliable serum biomarkers to aid in the diagnosis of peripheral arterial disease (PAD). We hypothesized that circulating Fatty Acid Synthase (cFAS) can be an independent diagnostic biomarker for PAD. Serum cFAS and demographics were compared for patients with and without PAD or CLTI. Patients with PAD or CLTI had significantly higher serum cFAS content. We observed optimal cutoffs for cFAS in distinguishing between individuals with and without PAD or CLTI. Our study demonstrates that cFAS is an independent serum-based diagnostic biomarker for PAD, can distinguish between patients with PAD versus CLTI, and may predict disease severity.

## 1) BACKGROUND

Peripheral arterial disease (PAD) affects over 230 million people worldwide, including 8.5 million in the United States alone ^1–3^. Managing PAD in the U.S. imposes an annual cost exceeding $20 billion, placing a substantial burden on the healthcare system ^4,5^. Individuals with PAD face an elevated risk of poly-vascular complications, including myocardial infarction and stroke, which lead to increased morbidity and mortality ^1,2,6^. The majority of individuals with PAD are asymptomatic and often only receive a diagnosis after progressing to disabling symptoms ^7,8^. Given that only 11% of patients present with ‘classical’ claudication symptoms ^7^, and up to 25% may advance to severe disease stages, such as chronic limb-threatening ischemia (CLTI) ^3^, improved early detection strategies are crucial. Despite the strong association between PAD and high-risk atherosclerotic profiles, individuals with undiagnosed PAD are less likely to receive risk factor modification and preventive therapies ^7,8^. Evidence is mounting that enhanced early diagnosis of PAD in high-risk populations can mitigate disease progression and related complications ^9–11^.

Current clinical guidelines emphasize the importance of assessing atherosclerotic cardiovascular disease (ASCVD) risk using serum low-density lipoprotein (LDL) levels and tools like the ASCVD 10-year risk calculator to estimate patient cardiovascular risk profiles ^9^. While LDL correlates with cardiac disease progression, studies have yet to establish its association with PAD severity, and it has not been shown that statin therapy can reduce the risk of major lower-extremity amputations resulting from PAD or CLTI ^12–14^. The Ankle Brachial Index (ABI) remains the most commonly used tool for PAD diagnosis ^15,16^; however, both the USPSTF and 2016 AHA/ACC guidelines highlight insufficient evidence supporting the reliability of ABIs as a screening test for asymptomatic patients ^2,17^. Moreover, ABI results can be falsely elevated in populations such as those with diabetes or end-stage renal disease, further limiting its accuracy ^7,15,18,19^. Despite these limitations, ABI remains the primary screening tool for PAD, underscoring the need for more reliable and accessible diagnostic options ^7,15,16^.

Fatty Acid Synthase (FAS) is a multifunctional enzyme that catalyzes the biosynthesis of fatty acids (FAs), which are vital for cellular membrane integrity and secondary signaling functions across nearly all life forms ^20,21^. Recently, a circulating serum form of FAS (cFAS) was identified in individuals with an elevated atherosclerotic disease burden ^22,23^. In an initial cohort of patients with chronic limb-threatening ischemia (CLTI), cFAS emerged as an independent risk factor for disease severity, irrespective of diabetes status or smoking history ^23^. Further evidence suggests that cFAS is not merely a byproduct but may actively contribute to the pathogenesis of atherosclerosis ^24,25^. Given these observations, we investigated whether cFAS could serve as a sensitive serum biomarker for detecting PAD across varying levels of disease severity.

## 2) METHODS

### 2.1) Patient Cohort

We conducted a retrospective review of individuals who participated in the vascular biobank over a 9-year period (2014-2023). Participants were categorized as healthy controls without PAD, individuals with PAD, or individuals with CLTI, based on the ABI and Rutherford score ^3,26^. Patients were excluded if they had been included in prior published studies or had a history of stage 4 or 5 chronic kidney disease (CKD), alcohol abuse, or advanced liver disease (**Fig. 1**). Demographic data collected included age at the time of serum sampling, sex, body mass index (BMI), race/ethnicity, and smoking status (prior/current). Medical history, medication use, and laboratory values were obtained from clinical chart reviews. To calculate the Framingham Risk Score (FRS) ^27^, available patient demographics were entered into the 2018 Prevention Guidelines CV Risk Calculator ^9,28^. Patient values outside the calculator’s parameters were rounded to the nearest valid value (e.g., age <40 was rounded up to 40, and TG <130 was rounded up to 130).

**Figure 1:**
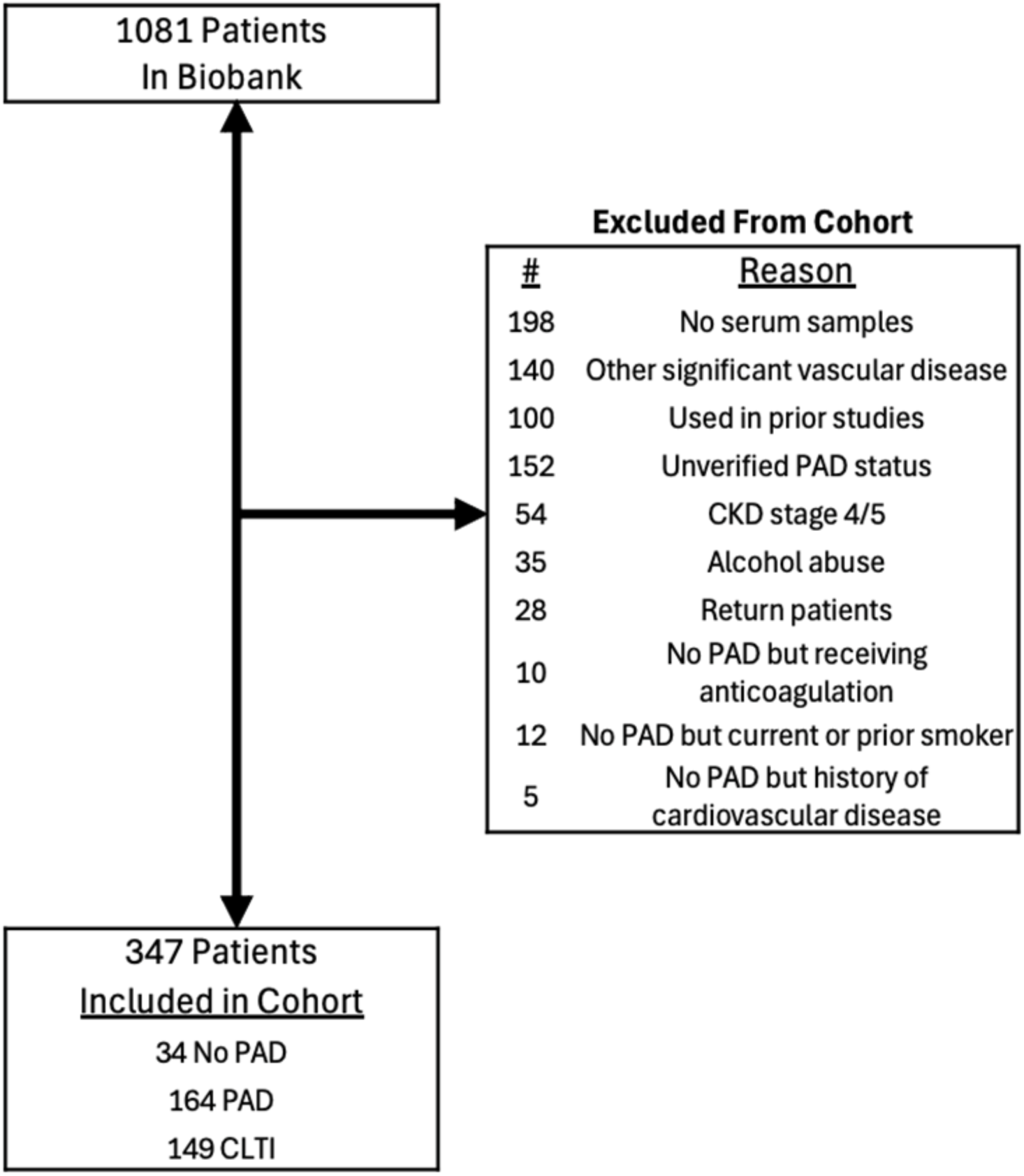
Flowchart of study patients. Patients were selected for the study from an initial pool of 1,081 patients in our vascular biobank. A total of 734 patients were excluded for reasons including lack of serum samples (n=198), presence of other significant vascular disease (n=140), prior inclusion in other studies (n=100), unverified PAD status (n=152), advanced chronic kidney disease (CKD stage 4/5; n=54), history of alcohol abuse (n=35), repeat patients (n=28), anticoagulation use without PAD (n=10), smoking history without PAD (n=12), and cardiovascular disease without PAD (n=5). The final cohort included 347 patients, categorized as 34 without PAD, 164 with PAD, and 149 with CLTI.

### 2.2) Blood Collection and Processing

Intravenous whole blood samples were collected from consenting individuals who were fasting for at least 6 hours prior to a planned elective surgery. As previously described, whole blood samples were collected in red-topped and green-topped vacutainer tubes, and immediately processed in the laboratory with centrifugation to isolate serum and plasma components ^29^. Serum and plasma were then aliquoted into 100 μL fractions and stored at -80° C for future analytical use.

### 2.3) Serum and Plasma Analysis

As previously described, serum aliquots were used to measure cFAS with a commercially available ELISA (Aviva Systems Biology, San Diego, CA) ^22–24^. To account for variations in the duration of fasting prior to surgery, total protein levels were determined via Bradford assay and used to normalize serum cFAS concentrations. Plasma samples were analyzed at the Washington University Diabetes Research Center (DRC) Core Laboratory for Clinical Studies (CLCS) for measurement of total cholesterol (TC), triglyceride (TG), direct high-density lipoprotein (HDL), and LDL.

### 2.4) Statistical Analysis

Receiver-operating characteristic (ROC) analysis is a statistical tool used to assess the diagnostic performance of a biomarker by calculating the area under the curve (AUC), which represents the measure of the ability of the test to correctly classify individuals as having or not having a disease. ROC analyses were conducted to determine the optimal cutoff points for circulating cFAS levels in distinguishing between different groups of patients: those who are normal, those with PAD, and those with CLTI. Multivariable regressions were built to measure the independent effect of cFAS threshold on patient group classifications. Models with best AIC (Akaike Information Criterion) and BIC (Bayesian Information Criterion) were selected. Chi-square test or Fisher’s exact test was utilized for categorical variables. Continuous variables were analyzed with two-sided t-test if normally distributed and Mann–Whitney U test if non-Gaussian. Categorical variables are represented as a number (percentage). All statistical analyses were performed using the R software (version 4.3.1).

### 2.5) Ethics

This study was approved by the Washington University in St. Louis School of Medicine institutional review board (IRB). All patients included in this study provided written informed consent to participate in a prospective maintained institutional vascular surgery registry and serum biobank.

## 3) RESULTS

### 3.1) Differences between Study Groups

Of the 1081 patients reviewed in the vascular biobank, a total of 347 patients met the inclusion criteria (**Fig 1**). Of these, 34 (9.8%) were healthy controls with no PAD, 164 (47.3%) had PAD, and 149 (42.9%) had CLTI (**Fig 2**). The normal group had a significantly younger mean age of 26.2 years (±5.4) compared to the PAD group at 64.8 years (±10.2) and the CLTI group at 63.4 years (±10.6) (p<0.001). The Body Mass Index (BMI) was numerically higher in the PAD group (28.6 ±5.9) and CLTI group (27.4 ±6.5) compared to the No PAD group (26.7 ±5.8, p=0.120). Gender distribution revealed a higher proportion of males in the PAD (64.6%) and CLTI (67.8%) groups compared to the No PAD group (41.2%, p=0.016). Most notably, cFAS levels were significantly elevated in the PAD group (560 pg/mg ± 680) and the CLTI group (610 pg/mg ± 740) compared to the No PAD group (210 pg/mg ± 280, p=0.007; **Fig 2, Tables 1-3**).

**Figure 2:**
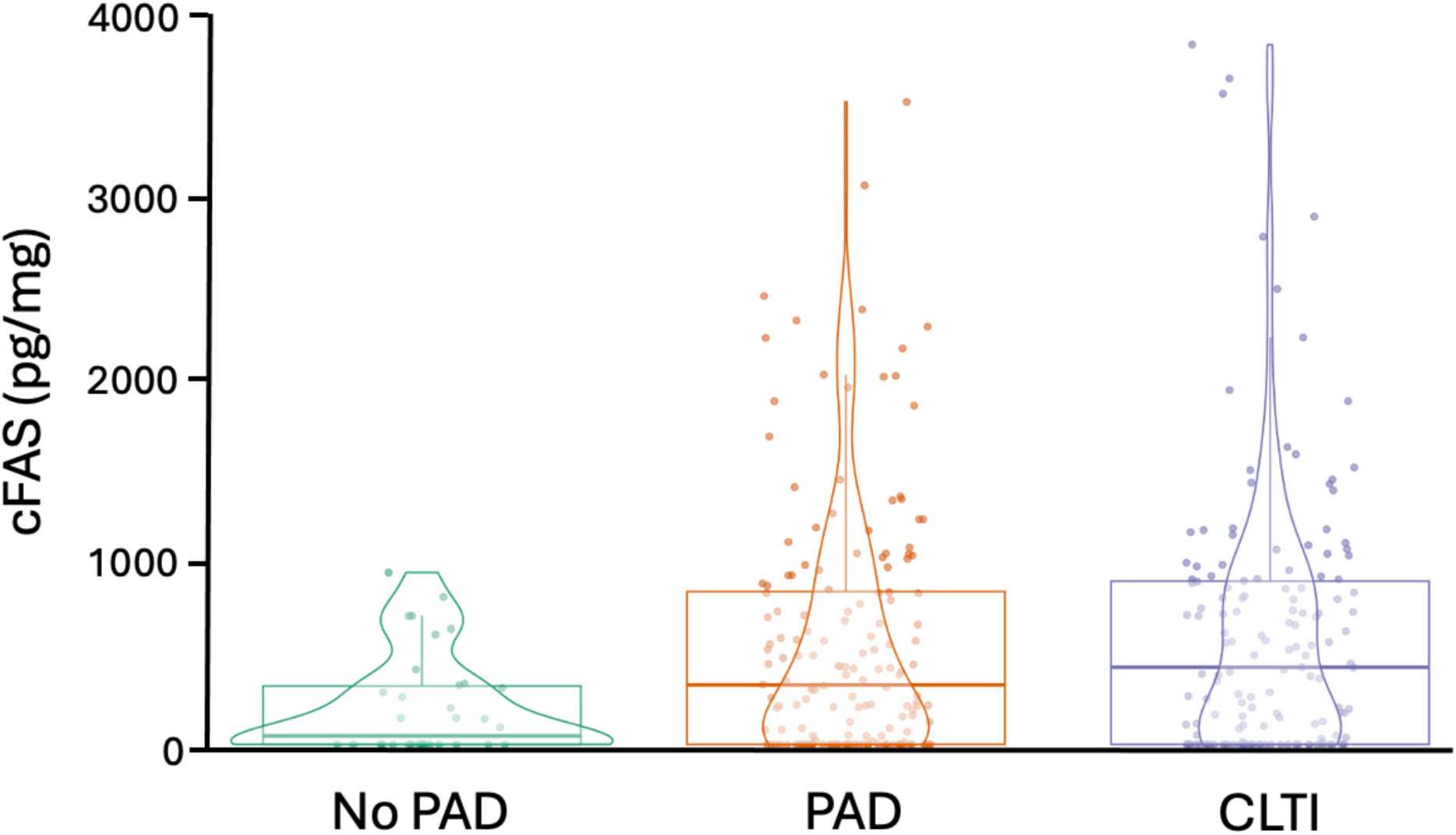
Violin plot displaying serum cFAS levels across three patient groups: No PAD, PAD, and CLTI. The distribution and density of serum cFAS levels are shown for each group, with individual data points overlaid. The median and interquartile ranges are represented within the box plots embedded in each violin. Serum cFAS levels are markedly higher in the PAD and CLTI groups compared to the No PAD group, with the highest levels observed in patients with CLTI, indicating a potential correlation between cFAS levels and PAD severity.

**Table 1.** Cohort Characteristics and Demographics, Individual Group Comparison

| Patient Characteristics | No PAD<br>(N=34) | PAD<br>(N=164) | CLTI<br>(N=149) | p-value |
| --- | --- | --- | --- | --- |
| Age (mean ± SD) | 26.2 (5.4) | 64.8 (10.2) | 63.4 (10.6) | **<0.001** |
| Body Mass Index (BMI) (mean ± SD) | 26.7 (5.8) | 28.6 (5.9) | 27.4 (6.5) | 0.1200 |
| Gender (Male %) | 14 (41.2) | 106 (64.6) | 101 (67.8) | **0.0160** |
| cFAS (ng/mg) (mean ± SD) | 0.2 (0.3) | 0.6 (0.7) | 0.6 (0.7) | **0.0070** |
| Creatinine (mg/dL) (mean ± SD) | 0.9 (0.2) | 1.0 (0.3) | 1.0 (0.3) | **0.0340** |
| Estimated Glomerular Filtration Rate (eGFR) (mean ± SD) | 105.2 (14.8) | 77.8 (21.1) | 79.8 (22.0) | **<0.001** |
| Triglycerides (mg/dL) (mean ± SD) | 87.1 (41.7) | 151.2 (99.3) | 148.9 (97.1) | **0.0010** |
| Total Cholesterol (mg/dL) (mean ± SD) | 163.4 (38.5) | 148.0 (45.2) | 138.9 (42.3) | **0.0080** |
| Direct HDL Cholesterol (mg/dL) (mean ± SD) | 48.9 (11.1) | 41.4 (13.5) | 38.4 (13.4) | **<0.001** |
| Direct LDL Cholesterol (mg/dL) (mean ± SD) | 99.3 (32.2) | 82.1 (35.9) | 77.4 (34.6) | **0.0050** |
| Framingham Risk Score (mean ± SD) | 0.01 (0.01) | 0.3 (0.2) | 0.2 (0.1) | **<0.001** |
| Carotid Artery Disease, N (%) | 0 (0.0) | 37 (22.6) | 16 (10.7) |  |
| Daily Aspirin Use, N (%) | 0 (0.0) | 119 (72.6) | 105 (70.5) | **<0.001** |
| Statin Use, N (%) | 0 (0.0) | 121 (73.8) | 111 (74.5) | **<0.001** |
| Insulin Use, N (%) | 0 (0.0) | 28 (17.1) | 32 (21.5) | **0.003** |
| Smoking or Tobacco Use Status, N (%) |  |  |  |  |
| Never used tobacco product (e.g., < 100 cigarettes in lifetime) | 32 (94.1) | 13 (7.9) | 14 (9.4) | **<0.001** |
| Current tobacco product user or quit within the past year | 0 (0.0) | 78 (47.6) | 76 (51.0) |  |
| Former tobacco product user (quit > 1 year ago) | 2 (5.9) | 73 (44.5) | 59 (39.6) |  |
| Diabetes Mellitus (DM), N (%) | 0 (0.0) | 76 (46.3) | 49 (32.9) | **<0.001** |
| History of Myocardial Infarction, N (%) | 0 (0.0) | 35 (21.3) | 30 (20.1) | **0.003** |
| History of Coronary Artery Disease (CAD), N (%) | 0 (0.0) | 80 (48.8) | 61 (40.9) | **<0.001** |
| History of Hypertension, N (%) | 0 (0.0) | 139 (84.8) | 118 (79.2) | **<0.001** |
| History of Cerebrovascular Disease, N (%) | 0 (0.0) | 58 (35.4) | 33 (22.1) | **<0.001** |

**Table 2.**
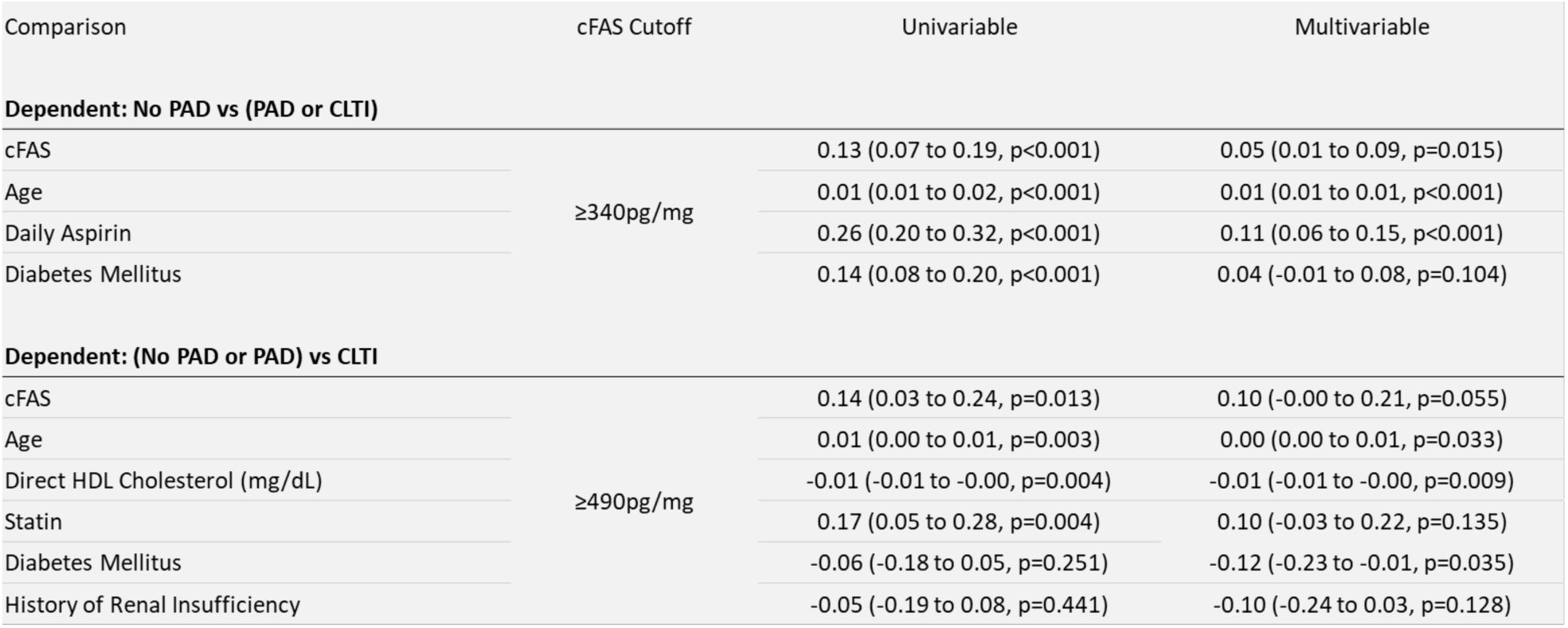
Univariate and Multivariate Analysis

Other notable findings observed between groups includes differences in total cholesterol (No PAD 163.4 ± 38.5 vs PAD 148.0 ± 45.2 vs CLTI 138.9 ± 42.3, p=0.008), triglycerides (No PAD 87.1 ± 41.7 vs PAD 151.2 ± 99.3 vs CLTI 148.9 ± 97.1, p=0.001), LDL (No PAD 99.3 ± 32.2 vs PAD 82.1 ± 35.9 vs CLTI 77.4 ± 34.6, p=0.005), HDL (No PAD 48.9 ± 11.1 vs PAD 41.4 ± 13.5 vs CLTI 38.4 ± 13.4, p<0.001), and Framingham Risk Score (No PAD 1% ± 1% vs PAD 25% ± 16% vs CLTI 21% ± 13%, p<0.001).

### 3.2) cFAS Differences between Study Groups

ROC analysis identified two critical cutoff points for serum cFAS levels. A serum cFAS level of ≥340 pg/mg was determined to be the optimal threshold for differentiating individuals without PAD from those with either PAD or CLTI. At this threshold, the ROC curve demonstrated a good AUC, with a true positive rate (TPR) of 52.4% and a false positive rate (FPR) of 20.06% for PAD or CLTI (**Fig. 3**). For distinguishing CLTI specifically, a higher cFAS cutoff of ≥490 pg/mg was found to be optimal, yielding a TPR of 49.0% and an FPR of 35.9% (**Fig. 4**).

**Figure 3:**
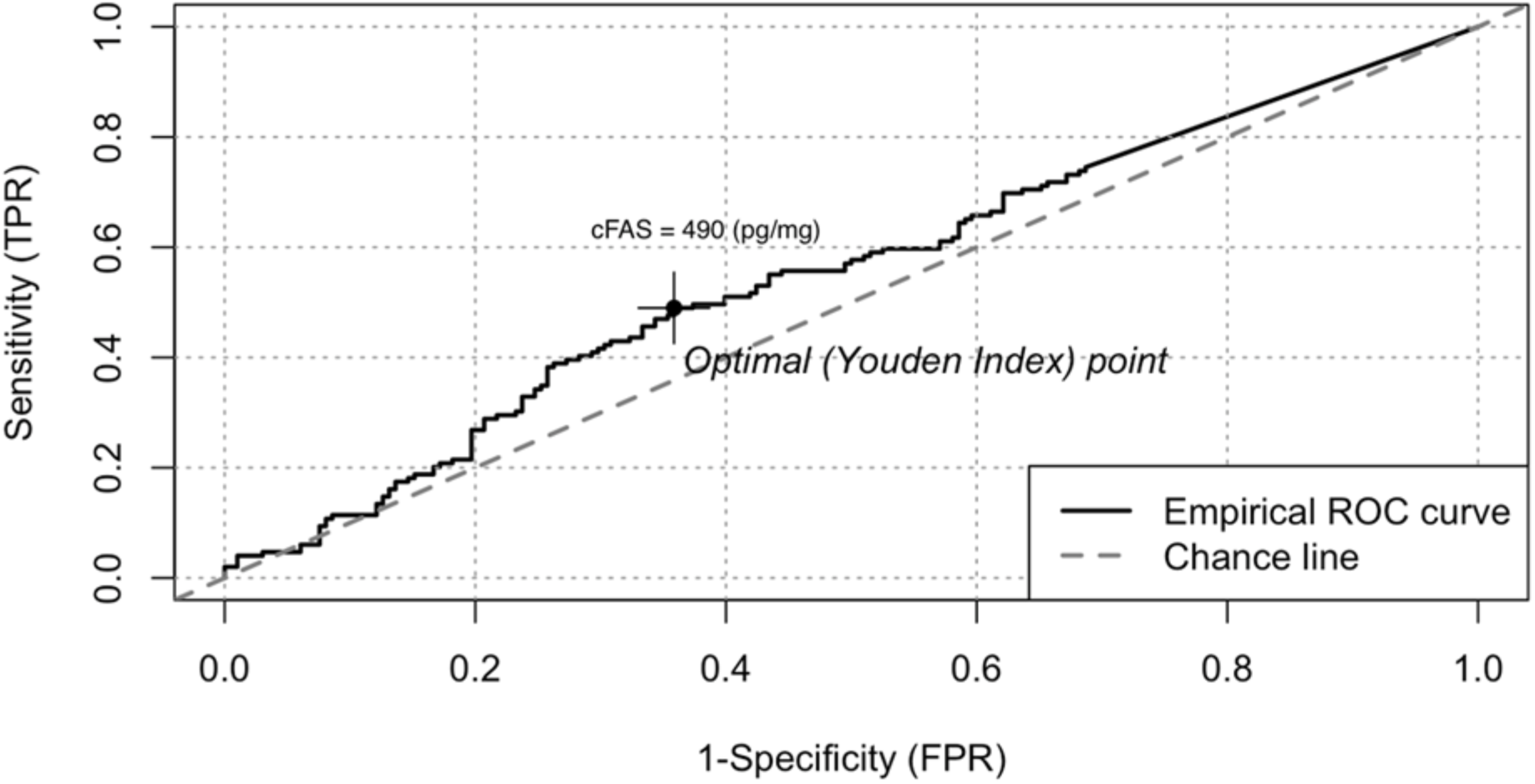
ROC curve illustrating the diagnostic performance of serum cFAS levels for distinguishing PAD from No PAD. The empirical ROC curve (solid line) shows sensitivity (True Positive Rate) versus 1-specificity (False Positive Rate) for various cFAS thresholds. The optimal cutoff point, determined using the Youden Index, is marked at cFAS = 0.34 ng/mg, which maximizes sensitivity and specificity. The dashed line represents the chance line, indicating random classification performance. This optimal cutoff highlights the diagnostic potential of cFAS for PAD detection.

**Figure 4:**
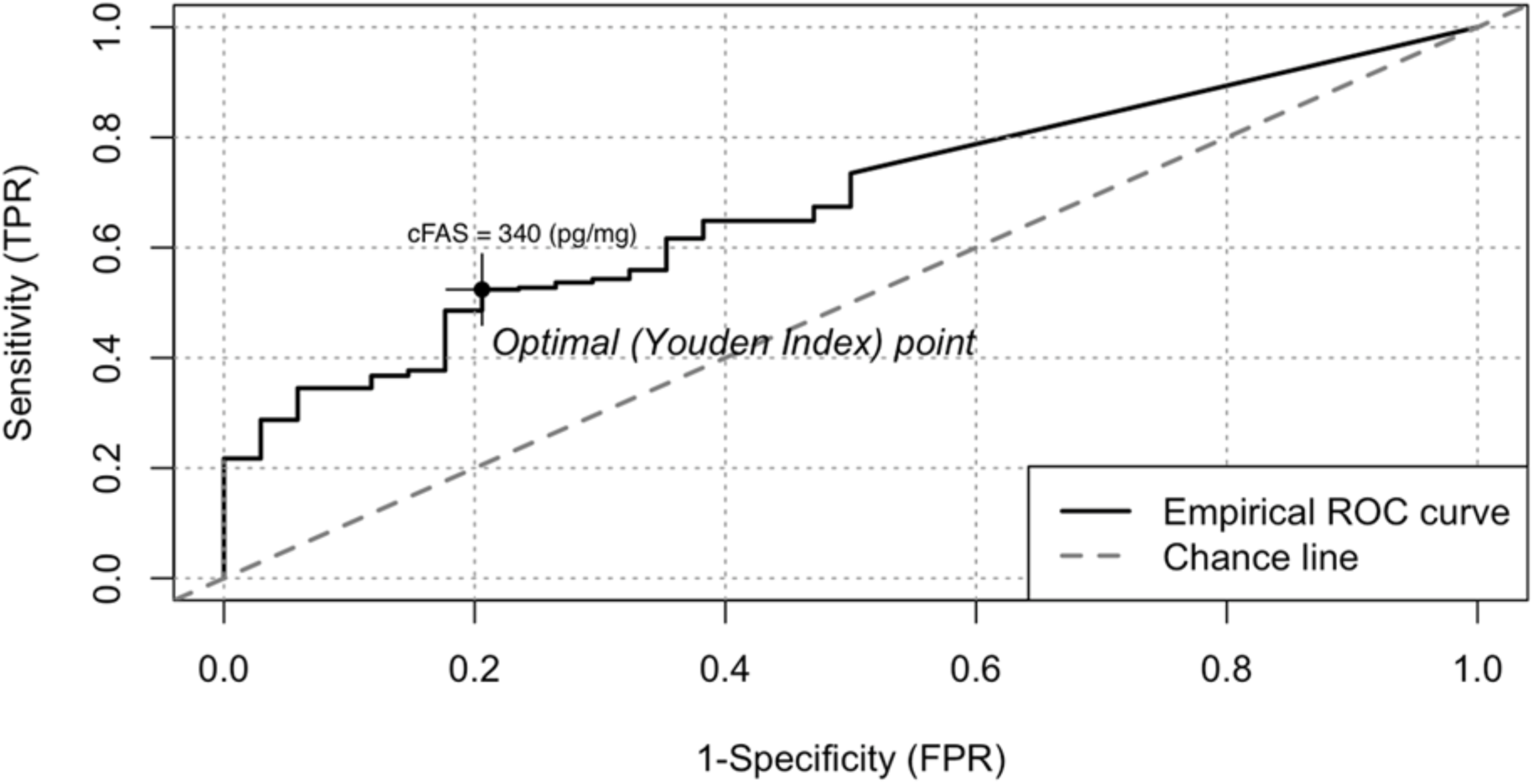
ROC curve showing the diagnostic performance of serum cFAS levels for distinguishing CLTI from No PAD or PAD. The empirical ROC curve (solid line) plots sensitivity (True Positive Rate) versus 1-specificity (False Positive Rate) across various cFAS thresholds. The optimal cutoff point, determined by the Youden Index, is indicated at cFAS = 0.49 ng/mg, maximizing sensitivity and specificity for CLTI detection. The dashed line represents the chance line, illustrating random classification. This cutoff underscores the potential of cFAS as a marker for advanced PAD stages such as CLTI.

Univariable analysis comparing individuals without PAD to those with PAD or CLTI indicated that a cFAS level of ≥340 pg/mg was associated with significantly higher odds of having PAD or CLTI, with an odds ratio (OR) of 0.13 (95% CI: 0.07 to 0.19, p<0.001). This finding suggests that elevated cFAS levels strongly correlate with the presence of PAD or CLTI. In a multivariable model adjusted for factors including age, diabetes status, HDL cholesterol, renal insufficiency, and statin and aspirin use, the association remained significant with an OR of 0.05 (95% CI: 0.01 to 0.09, p=0.015), reinforcing the independent relationship between elevated cFAS levels and PAD or CLTI.

In comparisons of individuals with no PAD or PAD, versus those with CLTI, the univariable model showed that a cFAS level of ≥490 pg/mg was associated with an odds ratio (OR) of 0.14 (95% CI: 0.03 to 0.24, p=0.013). This suggested that individuals with cFAS levels above this threshold are more likely to have CLTI than those without CLTI. In the multivariable model, adjusting for relevant factors, the adjusted OR was 0.10 (95% CI: -0.00 to 0.21, p=0.055), indicating a trend towards significance and suggesting that elevated cFAS levels may also independently correlate with CLTI (**Table 4**).

## 4) DISCUSSION

Our study evaluated whether serum cFAS could serve as a biomarker for PAD across varying levels of disease severity. We investigated the association between cFAS levels and PAD incidence, while adjusting for confounding factors such as age, sex, and comorbidities such as diabetes and smoking. ROC analysis revealed that serum cFAS levels were significantly associated with the presence of PAD and CLTI, highlighting its potential utility as a diagnostic marker. In our cohort of 347 tested patients, cFAS could identify PAD or CLTI (cutoff ≥340 pg/mg) with 52.4% sensitivity and 79.9% specificity. For identifying CLTI alone, a higher cutoff (≥490 pg/mg) demonstrated a sensitivity of 49% and specificity of 64.1%.

Recent studies support the association between serum cFAS levels and peripheral arterial disease beyond the coronary arteries ^22–24^. For instance, elevated serum cFAS has been observed in patients with carotid artery stenosis, particularly those with concomitant diabetes ^22^. Immunoprecipitation studies have demonstrated that the 275 kDa cFAS protein associates with Apolipoprotein B (ApoB), the primary apolipoprotein in LDL particles ^22^. Conditional knockdown of the *Fasn* gene in the liver significantly reduces serum cFAS levels, as seen in *Fasn^fl/fl^ Apoe^-/-^* mice, which also show reduced atherosclerotic plaque formation when maintained on a high-fat diet ^24^. In humans, elevated serum cFAS has been linked with higher FAS and saturated fatty acid content in the peripheral arteries, contributing to macrophage foam cell formation and atherosclerosis progression ^23,24^. These findings suggest that serum cFAS may serve as an indicator of atherosclerotic disease severity. In our current study we further demonstrate that cFAS levels are elevated in patients with confirmed PAD and reach the highest levels in those with CLTI.

This study deliberately excluded serum samples from patients included in previous publications to provide a new and independent assessment of cFAS as a marker for PAD ^22,23^. In our multivariable model, cFAS maintained its association with PAD and CLTI independent of other traditional vascular disease risk factors, building on earlier pilot studies and suggesting that cFAS may predict PAD disease risk better than LDL or ABI alone ^23^. Other studies have proposed that markers of fatty acid synthesis, such as saturated fatty acids, actively contribute to atherosclerotic disease progression beyond simple LDL risk stratification ^25,30,31^. Although the association between cFAS and PAD severity did not reach statistical significance at the higher threshold for CLTI, the observed trend suggests that cFAS could be valuable for identifying advanced disease stages in larger studies. This aligns with evidence indicating that CLTI encompasses a spectrum of end-stage PAD complications, including rest pain, non-healing wounds, tissue necrosis, and gangrene ^2,3^. Due to the limitations of our sample size, we did not stratify CLTI cases beyond Rutherford Class and did not base our assessments on anatomical disease severity or differentiate between atherosclerotic or thrombotic occlusions. Future studies should address the capacity of cFAS to provide a more nuanced diagnostic signal in patients with varying CLTI severities due to atherosclerosis.

The 2019 ASCVD guidelines reinforced LDL as a primary marker for cardiovascular risk assessment, largely based on studies showing reduced cardiovascular events in patients with lower LDL levels or those on statin therapy ^9^. Initially developed by the American College of Cardiology (ACC) and American Heart Association (AHA) in 2013 ^13^, these guidelines have since expanded to include a broader range of vasculopathies, including PAD and its complications. In the absence of a PAD-specific biomarker, LDL and ABI are frequently used as surrogate indicators of disease risk ^15,27,32^. However, large-scale studies consistently show significant underdiagnosis of PAD, particularly among asymptomatic individuals and those with atypical symptoms ^7,8^. For example, both the REACH Registry and the PARTNERS study reported that conventional screening measures failed to identify PAD in approximately 25% to 50% of cases ^7,8^, underscoring the need for more reliable diagnostic modalities in current clinical practice.

Our study showed that LDL did not correlate with the presence of PAD or CLTI. In fact, total cholesterol and LDL levels were generally lower among patients with PAD and CLTI compared to those without PAD, likely due to the use of cholesterol-lowering medications such as statins. Current AHA/ACC guidelines recommend LDL as an indicator for atherosclerosis and a trigger for statin therapy, alongside ABI for PAD screening ^9^. However, large-scale randomized controlled trials demonstrating LDL as a predictor for PAD are limited, and some studies suggest that LDL may not be a strong determinant of cardiovascular risk in specific populations, such as women ^9,33–35^. Our study found that cFAS was equally diagnostic in men and women, with sex not significantly influencing cFAS levels.

All participants in this study underwent an ABI test as the gold standard for diagnosing PAD and CLTI. The ACC/AHA guidelines reference Feigelson et al., which rigorously assessed ABI for PAD screening, noting an ABI <0.8 had a 39% sensitivity and 70% specificity for detecting PAD ^18^. Other studies have similarly shown variability in ABI sensitivity, depending on whether the ABI is high or low, and affected by operator expertise ^15,27,32,36^. Due to these limitations, the USPSTF does not currently endorse ABI testing for PAD screening, and the ABI is not reimbursed by CMS for asymptomatic patients ^17^. In contrast, a serum biomarker like cFAS, which has comparable sensitivity and specificity and requires only a small volume blood sample, could be accessible to primary care providers and has the potential identify patients with PAD.

We acknowledge several limitations of this study. Our patient cohort included individuals with varying medical histories and potential confounding variables that may have influenced the outcomes. Additionally, the negative control samples relied on self-reported patient histories, introducing potential recall bias. Our sample was also limited in capturing truly asymptomatic PAD cases, as our institutional biobank predominantly includes patients scheduled for surgical intervention. Although we included a No PAD control group, these individuals were generally younger and healthy except for thoracic outlet syndrome, which limits the generalizability of our findings. To enhance the analytical validity of cFAS as a biomarker, it will be essential to develop a more robust and rapid serum cFAS test. While incorporating factors such as age, gender, smoking status, and diabetes into our multivariable model was beneficial, it is possible that other confounding variables were not adequately represented in our ROC analyses.

### Clinical Perspective

- No laboratory blood tests is available to help in identifying patients with peripheral arterial disease (PAD)
- Fatty acid synthase (FAS) plays an important role in atheroprogression
- Serum circulating FAS (cFAS) is elevated in patients with chronic limb threatening ischemia (CLTI)

## Data Availability

All data produced in the present study are available upon reasonable request to the authors

## ACKNOWLEDGMENTS

None

---

**Supplemental Table 1.** Cohort Characteristics and Demographics, Comparison to Normal

| Patient Characteristics | No PAD<br>(N=34) | PAD and CLTI<br>(N=313) | p-value |
| --- | --- | --- | --- |
| Age (mean ± SD) | 26.2 (5.4) | 64.1 (10.4) | **<0.001** |
| Body Mass Index (BMI) (mean ± SD) | 26.7 (5.8) | 28.0 (6.2) | 0.2240 |
| Gender (Male %) | 14 (41.2) | 207 (66.1) | **0.0080** |
| cFAS (ng/mg) (mean ± SD) | 0.2 (0.3) | 0.6 (0.7) | **0.0020** |
| Creatinine (mg/dL) (mean ± SD) | 0.9 (0.2) | 1.0 (0.3) | **0.0110** |
| Estimated Glomerular Filtration Rate (eGFR) (mean ± SD) | 105.2 (14.8) | 78.7 (21.5) | **<0.001** |
| Triglycerides (mg/dL) (mean ± SD) | 87.1 (41.7) | 150.2 (98.1) | **0.0130** |
| Total Cholesterol (mg/dL) (mean ± SD) | 163.4 (38.5) | 143.7 (44.0) | <0.001 |
| Direct HDL Cholesterol (mg/dL) (mean ± SD) | 48.9 (11.1) | 40.0 (13.5) | **0.0020** |
| Direct LDL Cholesterol (mg/dL) (mean ± SD) | 99.3 (32.2) | 79.9 (35.3) | <0.001 |
| Framingham Risk Score (mean ± SD) | 0.01 (0.01) | 0.2 (0.2) | **<0.001** |
| Carotid Artery Disease, N (%) | 0 (0.0) | 53 (16.9) | **0.0080** |
| Daily Aspirin Use, N (%) | 0 (0.0) | 224 (71.6) | **<0.001** |
| Statin Use, N (%) | 0 (0.0) | 232 (74.1) | **<0.001** |
| Insulin Use, N (%) | 0 (0.0) | 60 (19.2) | **0.002** |
| Smoking or Tobacco Use Status, N (%) |  |  |  |
| Never used tobacco product (e.g., < 100 cigarettes in lifetime) | 32 (94.1) | 27 (8.6) | **<0.001** |
| Current tobacco product user or quit within the past year | 0 (0.0) | 154 (49.2) |  |
| Former tobacco product user (quit > 1 year ago) | 2 (5.9) | 132 (42.2) |  |
| Diabetes Mellitus (DM), N (%) | 0 (0.0) | 125 (39.9) | **<0.001** |
| History of Myocardial Infarction, N (%) | 0 (0.0) | 65 (20.8) | **0.001** |
| History of Coronary Artery Disease (CAD), N (%) | 0 (0.0) | 141 (45.0) | **<0.001** |
| History of Hypertension, N (%) | 0 (0.0) | 257 (82.1) | **<0.001** |
| History of Cerebrovascular Disease, N (%) | 0 (0.0) | 91 (29.1) | **<0.001** |

**Supplemental Table 2.**
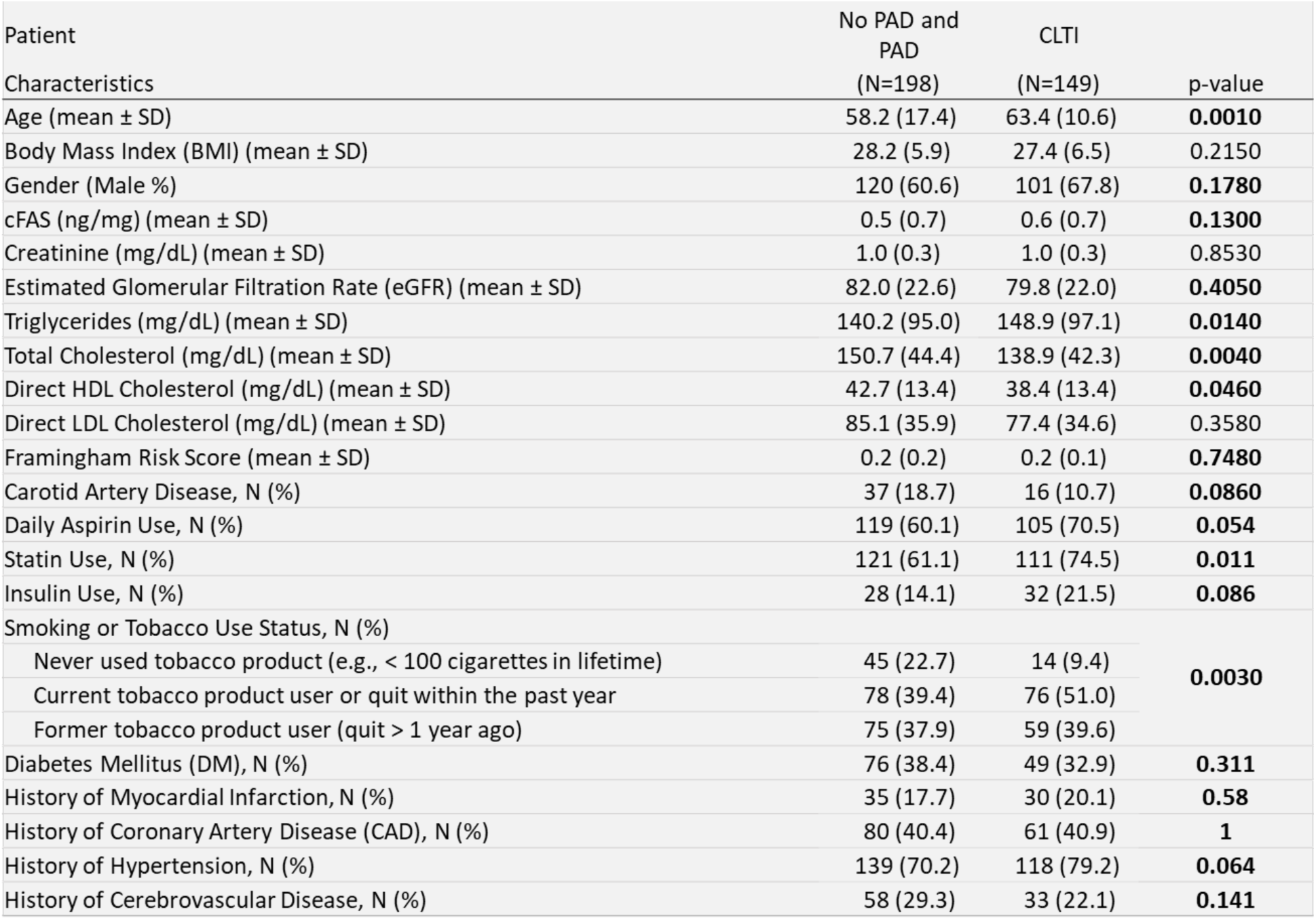
Cohort Characteristics and Demographics, Comparison to CLTI

